# A systematic review and meta-analysis protocol on the association of malaria in pregnancy and adverse birth outcomes

**DOI:** 10.1101/2020.07.19.20157081

**Authors:** Vikas Yadav, Mohan Bairwa, Deepti Dabar, Akhil Dhanesh Goel, Sarika Palepu, Ankur Joshi

**Author notes:** Corresponding author: Dr Vikas Yadav, Assistant Professor, Community Medicine, Atal Bihari Vajpayee Government Medical College, Vidisha, India.

## Abstract

**Introduction:** Malaria in pregnancy contributes to significant adverse birth outcomes. This study is aimed to quantify the relationship between malaria in pregnancy and occurrence of adverse birth outcomes, including preterm delivery, low birth weight, small for gestational age, miscarriages, and stillbirth.

**Methods and analysis:** Observational studies and Randomised controlled trials reporting data on selected birth outcomes separately for pregnancies, with or without malaria will be included. We will search for studies over various information sources and data extraction will be done from included studies. Pooled odds ratio (OR) will be calculated for each birth outcomes using fixed effect model or random effects models, based on the level of heterogeneity. Forest plot will be prepared with effect size (with 95 percent confidence interval) of each study and pooled effect size. The methodological quality will be assessed for included observational studies using the Newcastle-Ottawa scale (NOS). Cochrane Risk of Bias tool will be used to evaluate bias in randomised controlled trials. For publication bias, funnel plot will be prepared and assessed for asymmetry, along with Egger’s test.

**Discussion:** This study will provide an estimate of the risk of adverse birth outcomes in pregnancies with malaria. Results of this study will contribute towards planning effective service delivery in areas with a higher risk of malaria transmission.

**Ethics and dissemination:** The current study is a review of published literature, and it does not require ethical committee approval. Results of this review will be published in a peer-reviewed journal.

**PROSPERO registration number:** CRD42020153009

**Article summary:** *Strengths and limitations of this study:*

- To the best of reviewers’ knowledge, this will be a first-ever comprehensive review on the relationship between malaria in pregnancy and occurrence of most frequent adverse birth outcomes, i.e. preterm delivery, low birth weight, small for gestational age and miscarriages.
- The finding of this review will be very crucial for the governments of malaria-endemic countries.
- All the leading information sources will be included in this study to perform searches.
- The protocol is prepared in adherence to Preferred Reporting Items for Systematic Reviews and Meta-Analyses Protocols guidelines (PRISMA-P).
- A limitation of this study will be, various definition criteria used to define adverse birth outcomes among the included studies. Due to which, it will be challenging to interpret and extrapolate the findings of this study.

## Introduction

Worldwide, malaria is now the second leading cause of deaths due to infectious diseases after tuberculosis. In 2018, an estimated 228 million cases of malaria occurred worldwide with 405,000 deaths. Highest numbers were reported from the African countries followed by the South – East Asia region. Worldwide, the majority of the mortality burden (85%) of malaria is contributed by African countries and India. ^[1]^ Every year, 125 million women are at risk of Plasmodium falciparum or Plasmodium vivax malaria in pregnancy. ^[2]^

Although malaria affects all age groups and sexes, infection during pregnancy is a significant public health problem. It poses risks not only for the pregnant woman, but also for the foetus, and the new-born child. It is estimated that pregnant women are at three times higher risk of developing malaria than non-pregnant counterparts. ^[3]^ This may be due to the emergence of pregnancy-specific antigenic variants of Plasmodium falciparum that accumulate in the placenta.^[4-6]^ Immunological factors and hormonal changes in pregnancy also pre-dispose pregnant women to higher rates of malaria infection. ^[6]^

Many studies from areas with different malaria transmission patterns have investigated the consequences of malaria in pregnancy on both maternal health and birth outcomes. Maternal complications of malaria in pregnancy are anaemia, cerebral malaria and in severe cases, death. ^[7]^ Major complications among foetuses and new-borns, include, low birth weight, intrauterine growth retardation, abortion, stillbirth, premature delivery, and foetal death. ^[^8, 9^]^ Harmful effects on neonatal health are due to the accumulation of infected erythrocytes and their sequestration in the placental intervillous space, which leads to placental histopathological changes that trigger an exacerbated inflammatory response that is highly detrimental. The increased inflammatory response impairs the maternal-foetal interface and disrupts critical placental functions. ^[^6, 10, 11^]^ There is a paucity of reviews providing evidence on the relationship of malaria in pregnancy with adverse birth outcomes. However, Moore et al. (2017) ^[12]^ provided sufficient evidence about the relationship of malaria in pregnancy with stillbirth; there is a need for the evidence on the relationship of malaria in pregnancy with other adverse birth outcomes. In the current systematic review, we aim to assess the association of malaria in pregnancy with adverse birth outcomes, namely, preterm delivery, low birth weight, small for gestational age, and miscarriage. Stillbirth, another possible adverse outcome will be included in the systematic review, if there is any recent evidence, published after Moore et al. Thus, the current systematic review will help in fulfilling the knowledge gaps related to the effect of malaria in pregnancy on birth outcomes.

## Methods and analysis

We wrote this systematic review protocol according to the Preferred Reporting Items for Systematic Review and Meta-Analysis Protocols Guidelines (PRISMA-P). ^[13]^ The protocol has been registered on PROSPERO. ^[14]^

### Eligibility Criteria

The inclusion criteria developed using PECOS (Population, Exposure, Comparator, Outcomes, Study characteristics) framework ^[15]^ are as follows:

#### Study population (P)

Pregnant women, those who have been tested for malaria by PCR, light microscopy, rapid diagnostic test, or histology

#### Type of exposure (E)

Exposure of interest will be malaria infection (Plasmodium falciparum and/or Plasmodium vivax) in pregnancy.

#### Comparator (E)

Comparator group will comprise of pregnant women not suffering from malaria.

#### Outcomes (O)

Adverse birth outcomes, namely preterm delivery, low birth weight, small for gestational age, miscarriages and stillbirth, occurred to pregnant women.

Definition of birth outcomes: We will consider the definitions of adverse birth outcomes provided by authors of included articles. If authors do not provide definitions or there is some disparity, we will use these standard definitions: ^[16]^

- Preterm delivery: Birth before 37 completed weeks gestation.
- Low birth weight (LBW): Birth weight <2500g.
- Small for gestational age (SGA): Birth weight <10th centile for gestational age.
- Spontaneous abortion (miscarriage): A conceptus born after spontaneous labour without any signs of life before 24 completed weeks gestation.
- Stillbirth: Foetal death, after 24 completed weeks gestation and before delivery.

#### Study characteristics (S)

Eligible study designs will be observational studies including cohort, case-control, cross-sectional and selected randomised controlled trials (RCT). The RCTs, assessing the efficacy of a standard therapy drug (or drugs) on prevention of malaria (Prevention Trials) in pregnant women and providing data on any of the included birth outcomes in malaria-infected and non-infected pregnant women, will be included.

### Exclusion criteria

1. Case reports, case series, opinion, review articles and studies without comparison groups will be excluded.
2. Studies not providing data regarding any of the selected birth outcomes (preterm delivery, low birth weight, small for gestational age, miscarriages, and stillbirth) separately for malaria-infected and non-infected pregnant women, will also be excluded.

### Information sources

We will search Medline (via PubMed available at https://pubmed.gov/), Embase (https://www.embase.com), Scopus (https://www.scopus.com), Cochrane Library (https://www.cochranelibrary.com/) and The Malaria in Pregnancy Consortium Library (http://library.mip-consortium.org/) to identify studies reporting adverse pregnancy outcomes in women, who had malaria infection during the pregnancy.

Any date or language restrictions will not be applied in this review. The search strategy for this systematic review is developed based on the PECOS framework of the research question. The search terms will include, Study population (pregnant, prenatal, antenatal); Exposure of interest (malaria, placental malaria); and Outcomes of interest (preterm delivery, low birth weight, small for gestational age, miscarriages, and stillbirth). To develop a robust search strategy, we will use controlled descriptors (such as MeSH terms, Embase Emtree) and Boolean operators, in addition to identified keywords based on the PECOS framework. The full search strategy for PubMed is provided in Table 1.

**Table 1.**
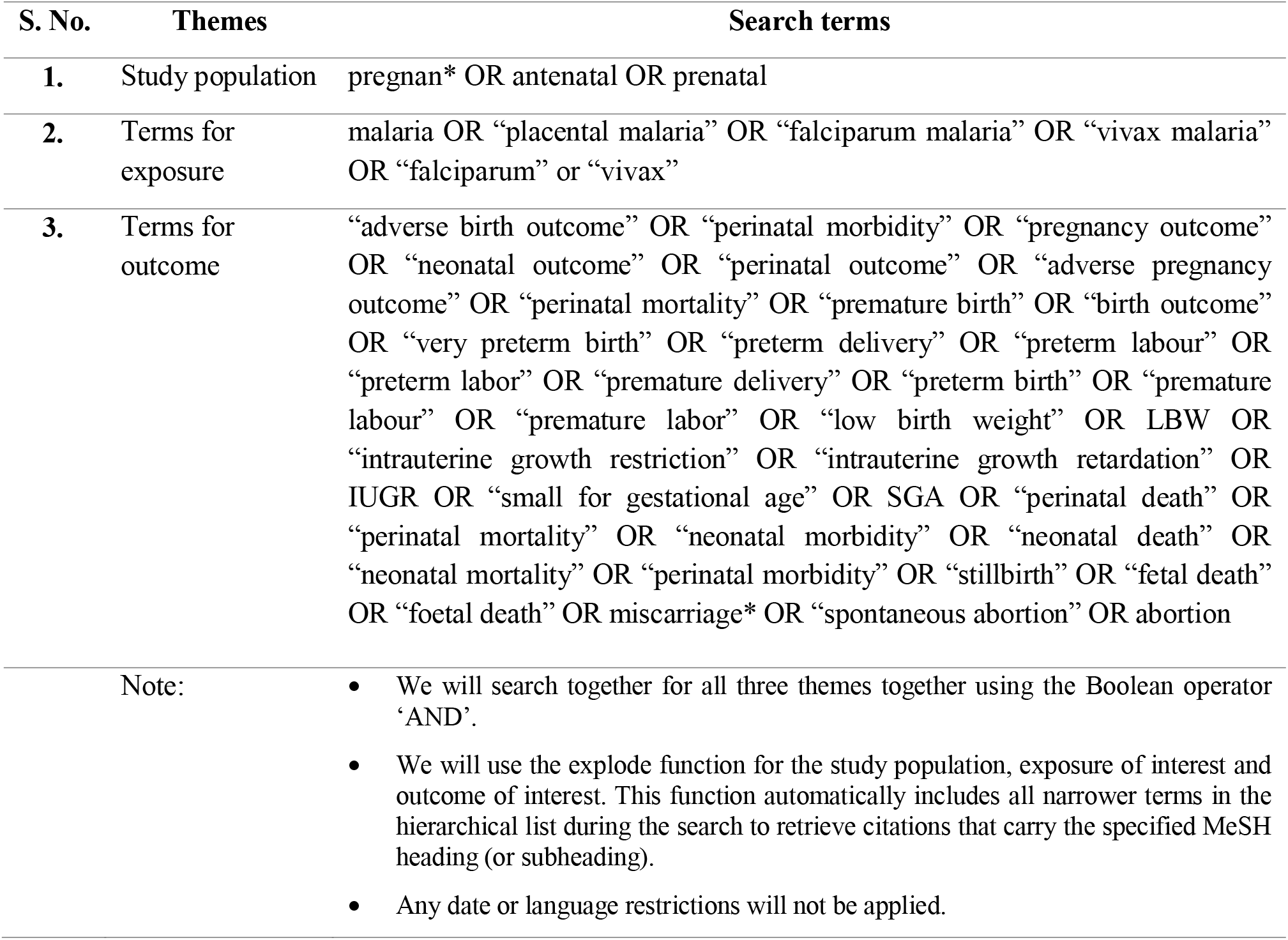
Search strategy for PubMed.

| S. No. | Themes | Search terms |
| --- | --- | --- |
| 1. | Study population | pregnan* OR antenatal OR prenatal |
| 2. | Terms for exposure | malaria OR “placental malaria” OR “falciparum malaria” OR “vivax malaria” OR “falciparum” or “vivax” |
| 3. | Terms for outcome | “adverse birth outcome” OR “perinatal morbidity” OR “pregnancy outcome” OR “neonatal outcome” OR “perinatal outcome” OR “adverse pregnancy outcome” OR “perinatal mortality” OR “premature birth” OR “birth outcome” OR “very preterm birth” OR “preterm delivery” OR “preterm labour” OR “preterm labor” OR “premature delivery” OR “preterm birth” OR “premature labour” OR “premature labor” OR “low birth weight” OR LBW OR “intrauterine growth restriction” OR “intrauterine growth retardation” OR IUGR OR “small for gestational age” OR SGA OR “perinatal death” OR “perinatal mortality” OR “neonatal morbidity” OR “neonatal death” OR “neonatal mortality” OR “perinatal morbidity” OR “stillbirth” OR “fetal death” OR “foetal death” OR miscarriage* OR “spontaneous abortion” OR abortion |
| Note: |  | • We will search together for all three themes together using the Boolean operator ‘AND’.<br>• We will use the explode function for the study population, exposure of interest and outcome of interest. This function automatically includes all narrower terms in the hierarchical list during the search to retrieve citations that carry the specified MeSH heading (or subheading).<br>• Any date or language restrictions will not be applied. |

We will supplement the database searches by screening the bibliographies of the previous systematic reviews and relevant original research articles. The references of all publications identified in the primary search will also be inspected. We will contact leading researchers in the field for non-captured published articles with current search strategies.

### Study selection process

Two reviewers (ADG and SP) will independently conduct searches on all information sources. All search results will be uploaded into Rayyan QCRI online Software (https://rayyan.qcri.org). This will help ensure a systematic and comprehensive search and selection process. A third reviewer (VY) will manage Rayyan QCRI software, who will identify and remove the duplicate citations and ensure independent review of titles and abstracts by blinding the decisions of both reviewers.

The third reviewer will also identify the discrepancies between the two reviewers and discuss them, for making consensus to select the articles. Full-text copies of all selected studies will be obtained to find more details. Both reviewers (ADG and SP) will review the full text of articles and resolve the discrepancies by consensus. If needed, the arbitration will be done by the third reviewer (VY).

We will document the reasons for the exclusion of studies explored as full text. If any study is reported as multiple publications, all publications will be obtained. Though the study will be included only once in the review, data will be extracted from all the publications to collect maximum relevant data. The study inclusion process will be presented using the PRISMA flow chart. ^[17]^

The reference management software Mendeley Desktop (https://www.mendeley.com) for the Windows operating system will be used to store, organize, cite, and manage all the included articles.

### Data extraction process

Both reviewers (ADG and SP) will independently perform data extraction of relevant information regarding study details, population characteristics, malaria infection details, and birth outcomes. Any variation in the data abstraction will be resolved by consensus, and if required, the arbitration will be accomplished by other members of the review team (VY and DD). Authors of included studies will be contacted if missing or additional information will be required. Table 2 provided a comprehensive list of all the data items that will be extracted from selected studies.

**Table 2:** Comprehensive list of all data item to be extracted

| **Data items** | **Components of the data items** |
| --- | --- |
| **Study details** | authors, year of publication, study period, location, setting and study design |
| **Population characteristics** | number of pregnant women in malaria positive and malaria negative groups |
| **Infection details** | type of plasmodium species studied, malaria detection method, time of malaria detection (during pregnancy or delivery) |
| **Birth outcome** | frequency of each outcome in malaria positive and malaria negative groups: stillbirth, preterm delivery, low birth weight, small for gestational age and miscarriages; definitions of outcomes |

### Quality or risk of bias assessment of individual studies

Two reviewers (ADG and SP) will independently assess and score the quality of the selected observational studies according to the Newcastle-Ottawa scale (NOS).^[18]^ This scale is used for observational studies, where studies are scored between zero to nine stars for nine questions, that cover three areas: selection, comparability, and outcome. To quantifying the risk of bias of RCTs, two reviewers (ADG and SP) will independently assess studies using The Cochrane Collaboration’s tool for assessing the risk of bias in randomised trials. ^[19]^ It provides judgment as the study is having high, low, or unclear risk of bias. Inconsistencies in the findings will be resolved by discussion amongst both the reviewers (ADG and SP) and arbitration by the third reviewer if needed (VY).

### Data synthesis and analysis

Pooled OR will be calculated separately for each birth outcome (preterm delivery, low birth weight, small for gestational age, miscarriages, and stillbirth). We will use the appropriate model (fixed effect model or random effects models), based on the level of heterogeneity. Heterogeneity between studies will be examined using Cochran’s Q test and quantified using the I^2^ statistic. ^[20]^ Forest plot will be prepared, which will show the effect sizes and confidence intervals of individual studies, and pooled estimate of the effect size of all included studies.

Sensitivity analysis will be done by including higher-quality studies (observational studies) and studies with a lower risk of bias (RCTs). We will conduct subgroup analysis to investigate the possible source of heterogeneity based on the following variables: study design, country or region, malaria detection method, type of plasmodium species studied, time of birth outcome detection (during pregnancy or delivery). All the analysis will be conducted in updated versions of STATA software package (StataCorp LLC, TX) and R software using Metafor/Meta packages. ^[^21, 22^]^

### Publication bias

Publication bias will be evaluated by visual inspection of funnel plots and testing using Egger’s test, with p<0.1 considered indicative of statistically significant publication bias. ^[23]^

### Ethics and dissemination

This systematic review and meta-analysis comprise data from published literature only; hence, it does not require ethical approval. Results of this review will be published in a peer-review scientific journal.

## Discussion

Malaria remains a significant public health problem even decades after implementation of control programmes. The infection poses a dual burden in pregnant women by affecting the well-being of both mother and child. Malaria in pregnant women also poses an economic burden not only on the family but also on the health system of the country.^[11]^ Due to placental malaria infection, many pathological changes as inflammation, intervillous fibrin deposition, and infarction take place. These changes might be persistent until delivery and affect the maternal-foetal-placental unit, giving rise to adverse birth outcomes. ^[24]^ Also, women with infection in mid-pregnancy have higher chances of foetal growth retardation. ^[25]^

According to an estimate, in 2018, about 11 million pregnant women in moderate and high transmission regions of sub-Saharan African countries would have been exposed to malaria infection. Due to which, nearly 900,000 children were born with low birth weight. ^[1]^ However, to date, there is a lack of scientific evidence assessing the cumulative risk of adverse birth outcomes associated with this infection. This systematic review and meta-analysis strive to fill the gap and add new volumes to scientific evidence by quantifying the risk of a spectrum of adverse effects in the new-borns due to malaria in pregnancy.

The findings from this review will assist in the strategic planning of ante-natal care services for pregnant women residing in high transmission areas and are at risk of malaria infection. A multitude of morbidity and mortality due to various adverse effects would be assessed and outlined in terms of country, species of parasite, and duration of pregnancy in this review. Hence, sustainable scaling up and strengthening of essential health services can be planned in areas at higher risk of malaria transmission with light upon prevention and control of adverse outcomes.

The reviewers state that any modifications required in the protocol will be included with detailed description and justification during the publication of the review.

## Novelty

This systematic review, as far we know, is the first to assess the association between malaria in pregnancy and adverse birth outcomes, namely, preterm delivery, low birth weight, small for gestational age, and miscarriage.

## Patient consent for publication

Not required.

## Author contributions

The conception of the work: VY, MB, DD, ADG, SP, AJ. Designed the protocol and wrote the paper: VY, MB, DD, ADG, SP, AJ. Agreement to be accountable for all aspects of the work: VY, MB, DD, ADG, SP, AJ. All reviewers have read and approved the current version of the manuscript.

## Patient and public involvement

Patients or the public will not be involved in the design, or conduct, or reporting, or dissemination plans of this research.

## Data Availability

None

## Acknowledgements

None

## Data statement

None

